# Assessing the quality of nontraditional N95 filtering face-piece respirators available during the COVID-19 pandemic

**DOI:** 10.1101/2020.07.25.20161968

**Authors:** Deborah Plana, Enze Tian, Avilash K. Cramer, Helen Yang, Mary M. Carmack, Michael S. Sinha, Florence T. Bourgeois, Sherry H. Yu, Peter Masse, Jon Boyer, Minjune Kim, Jinhan Mo, Nicole R. LeBoeuf, Ju Li, Peter K. Sorger

## Abstract

**Background:** During the current COVID-19 pandemic, supply chains for Personal Protective Equipment (PPE) have been severely disrupted and many products, particularly surgical N95 filtering facepiece respirators (FFRs; “masks”) are in short supply. As a consequence, an Emergency Use Authorization (EUA) from the FDA has allowed importation of N95-type masks manufactured to international standards; these include KN95 masks from China and FFP2 masks from the European Union.

**Methods:** We conducted a survey of mask in the inventory of major academic medical centers in Boston, MA to determine provenance and manufacturer. We then assembled a simple apparatus for performing a necessary (but not sufficient) test of filtration performance and tested masks from the inventory; an accompanying website shows how to build and use the testing apparatus.

**Results:** Our survey showed that, seven months after the start of the COVID-19 pandemic, over 100 different makes and models of N95-type masks are in the inventory of local hospitals as opposed to 2-5 models under normal circumstances. A substantial number of unfamiliar masks are from unknown manufacturers. Many did not perform to accepted standards and are likely to be counterfeit. Due to the absence of publicly available information on mask suppliers in the FDA EUA and confusing or inconsistent labeling of KN95 masks, it is difficult to distinguish legitimate and counterfeit products.

**Conclusions:** Many of the FFR masks available for procurement during the COVID-19 pandemic do not provide levels of fit and filtration similar to those of N95 masks and are not acceptable for use in healthcare settings. Based on these results, and in consultation with occupational health officers, we make six recommendations for end users to assist in acquiring legitimate products. In particular, institutions should always assess masks from non-traditional supply chains by checking their markings and manufacturer information against data provided by NIOSH and the latest FDA EUA Appendix A. In the absence of verifiable information on the legitimacy of mask source, institutions should consider measuring mask fit and filtration directly. We also make suggestions for U.S and Chinese regulatory agencies with regard to labeling and public disclosure aimed at increase pandemic resilience.

## BACKGROUND

Filtering facepiece respirators (FFRs) such as N95 masks are the primary mode of respiratory protection for healthcare workers treating infectious agents that are airborne or transmissible via aerosols (1). As a result of the COVID-19 pandemic, demand for N95 masks and other personal protective equipment (PPE) has greatly outstripped supply, leading to widespread and persistent shortages. In the US, surgical N95 FFRs used in healthcare are regulated by the National Institute for Occupational Safety and Health (NIOSH), a part of the Centers for Disease Control and Prevention (CDC), and by the Food and Drug Administration (FDA) as described in US Code of Federal Regulations 42 CFR part 84 (2). Similar standards and enforcement mechanisms exist in other industrialized countries (3). Some FFRs with the filtering properties of healthcare N95 masks, including industrial N95 masks and elastomeric respirators, commonly have exhalation valves; such devices are traditionally not permitted for use healthcare settings because air exhaled through the valve is unfiltered, precluding the maintenance of a sterile field (4). Unfiltered exhalation through valves is also a possible avenue of disease transmission.

Very high demand for N95 respirators, coupled with disruption of medical supply chains, has led to a severe shortage of respiratory protection for healthcare workers during the 2020 COVID-19 pandemic (5). In February 2020, the FDA issued the first in a series of Emergency Use Authorizations (EUAs) relaxing regulations on N95 masks to help increase supply (6). These EUAs permitted the use in U.S. healthcare settings of masks manufactured for industrial use as well as of non-NIOSH approved masks meeting foreign standards functionally equivalent to those for N95 masks. As described in the EUAs “<u>Authorized Imported, Non-NIOSH Disposable Filtering Facepiece Respirators</u>” and “<u>Non-NIOSH Approved Disposable Filtering Facepiece Respirators Manufactured in China</u>” authorized masks include KN95 masks manufactured in China to the GB2626-2006 standard (7), FFP2 masks manufactured to European standard EN 149:2001, and masks manufactured in Australia, Brazil, Japan, Korea, and Mexico to other trusted performance standards (we refer to these collectively as N95-type masks) (8). As a practical matter, however, masks from China are the most common of these masks, reflecting China’s role as a leading producer of medical supplies.

Manufacturing N95-equivalent masks requires special fabrics and careful quality control and such masks must exhibit three essential functional properties: 1) the ability to filter out small particles (in the case of N95s, 95% of particles of the most penetrating aerosol size tested – typically down to 0.3 µm diameter); a tight fit to the face so that inhaled air is directed through the filter fabric and not around the side of the mask; and 3) low inhalation resistance so that a user does not experience difficulty breathing. Unfortunately data from the CDC (9) and other groups (10) has shown that, subsequent to the EUA permitting their use in US healthcare, some respirators manufactured overseas and labeled as N95, FFP2, or KN95 fail to perform as expected for filtration and fit. While this might be a consequence of manufacturing defects, it appears more likely that many of these non-performing respirators are counterfeit or claim adherence to standards that they never intended to meet (11). While a number of Chinese-brand respirators have performed well in quality testing, both unfamiliar products and counterfeits of known Chinese brands have been found in the US supply. Unfortunately, fraudulent packaging poor labelling practices can make it difficult to determine if a given respirator is genuine or not.

The first version of the FDA EUA on “Non-NIOSH Approved Disposable Filtering Facepiece Respirators Manufactured in China” (February 2020) included a list of authorized respirators and vendors in “Appendix A,” but no testing data was required from purported manufacturers to corroborate performance claims. As described below, it is hard to even identify the business addresses or websites of many of suppliers on Appendix A. Subsequent to the FDA EUA, the CDC noted a dramatic increase in counterfeit respirators with labeling that misrepresented such products as having been approved to NIOSH or equivalent foreign agencies (11). The CDC therefore began a program of performance testing and on May 7, 2020 the FDA substantially shortened Appendix A based on data demonstrating widespread inadequacies in filtration efficiency (12). The CDC continues to evaluate masks and to post photographs of the mask packaging for known counterfeit products, while the FDA continues to refine Appendix A. The most recent version Appendix A (issued on June 9, 2020) includes 144 FFR models from 86 manufacturers, down from 148 models and 68 manufacturers on July 13, 2020 (13).

In this paper we consider the problem of non-traditional N95-type masks from the perspective of an end user involved in healthcare, specifically large teaching hospitals affiliated with Harvard Medical School (HMS). For these users, one consequence of supply chain disruption and the initially permissive FDA EUA is that a large number of unfamiliar models of N95-type masks have become available (14), some of which come through irregular supply chains (14) or are donated and have unclear provenance. In a healthcare setting, fit testing masks on individual users is standard (e.g. using the 3M FT-30 qualitative fit test kit) (15), but hospitals are rarely if ever equipped to measure filtration efficiency (14). Such testing is usually performed by manufacturers either in-house or by commercial pre-certification laboratories on behalf of manufacturers. In the absence of such capabilities, end users are forced to evaluate masks from dozens of unknown manufacturers based on little or no information. In this paper, we attempt to assess the impact of these issues.

We inventoried masks on hand at HMS hospitals; attempted to match vendors and models to information in the FDA EUA Appendix A, and selected a subset for performance testing. We describe a simple filtration testing instrument assembled from commonly available components that can be used to determine if a mask is likely to meet performance standards: the instrument does not guarantee performance to N95-type standards but no legitimate mask should fail to exhibit at least 95% filtration efficiency using the apparatus we describe. We demonstrate multiple labeling and performance problems with non-traditional N95-style masks and formulate a set of recommendations to help guide healthcare organizations and other users in assessing mask donations and purchases. We also suggest ways in which the FDA and CDC can improve future EUAs.

## METHODS

Samples of N95-type masks 70 mm × 70 mm in size were cut from each mask and inserted into a circular acrylic air duct with an inner diameter of 50 mm (**Figure 1**). Either ambient particles or KCl aerosol particles were driven through the respirator filter using air flow to serve as a pollutant source. KCl aerosol was generated by a Collision Nebulizer (BGI Inc., USA) using 10 wt% KCl solution with the volume of free air set at 1 L/min. The concentration of 0.3-10 μm particles was determined using an optical particle counter (Aerotrak 9306, TSI Inc., USA). Concentrations were recorded twice at a one-minute intervals both upstream and downstream of the respirator filter, and the measurements then repeated once. The single-pass filtration efficiency *η* (d_p_) of particles with a size of *d*_*p*_ (μm) was calculated over an 8-minute test period as follows:

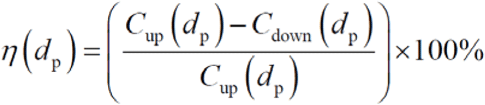

where *C*_*up*_ and *C*_*down*_ are the average particle counts (pcs) upstream and downstream of the filter, respectively. The pressure drop across the filter was measured by using a differential gauge. The air temperature was nominally *T* = 24 ± 1 °C and relative humidity 30 ± 20%; these values were not controlled but were measured along with air face velocity (which was typically in the range of 0.1 to 0.3 m/s) using a mini thermo-anemometer located at the air duct exhaust. Further information about the construction and use of this filter-testing instrument can be found at http://cleanmask.org.

**Figure 1.**
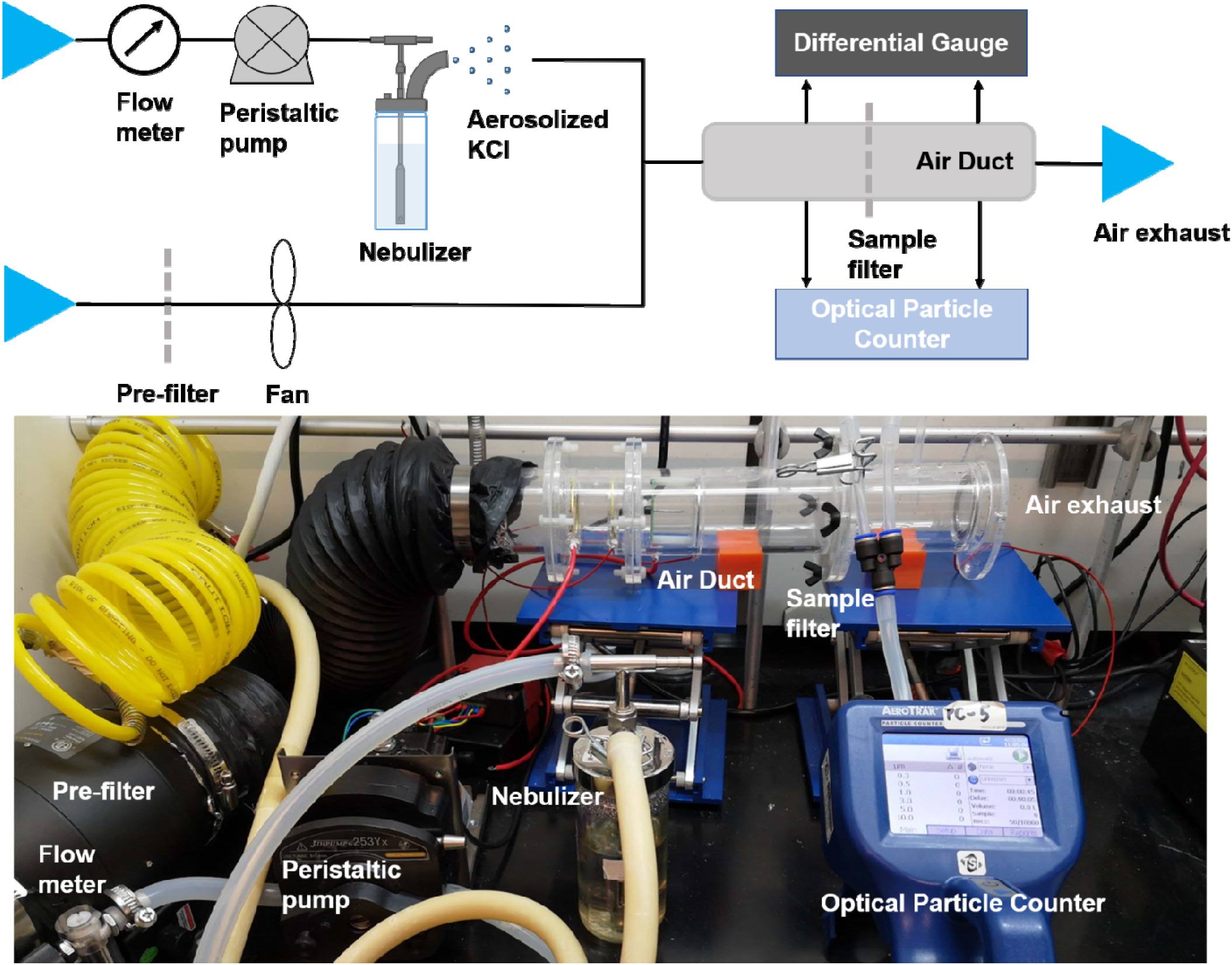
Apparatus assembled from common components and used to test FFRs in this study. Details of the fabrication and use of this device for testing the filtration ion efficiency of N95-type masks using ambient particles and KCl droplets can be found in supplementary materials and at http://cleanmask.org/setup. No legitimate FFR should demonstrate less than 95% filtration efficiency using this test, but testing performed with this apparatus is not sufficient to confirm performance to U.S., European, Chinese or other regulatory standards. Such testing involves a wider range of conditions, greater control over test conditions and a formal approach to quality assurance and calibration.

## RESULTS

### Qualitative examination of mask labeling helps identify legitimate respirators

To assess the diversity of the mask supply available for use during the COVID-19 pandemic, we inventoried models and makes of respirators, many donated, from HMS-affiliated medical centers in Boston, MA. We identified over 100 brands and models in the inventory. In contrast under, standard non-emergency conditions only two masks models, both from a traditional domestic manufacturer and provided via a familiar supply chain, would normally be in the inventory of the hospitals surveyed **(Table 1)**. A substantial number of the masks on hand originate from unknown vendors and appear to be of Chinese origin (based on the writing on the box). Many lacked even basic information such as manufacturer address or website, and respirator model numbers were generally lacking (**Figure 3C**). In some cases visual inspection revealed that masks in this inventory were similar in appearance or packaging to masks identified as counterfeit by the CDC and posted on their website (11). Moreover, a substantial number of masks listed multiple regulatory approvals from different countries. However, no mask claiming compliance to N95 standards should also claim compliance with KN95 or FFP2 standards because these are different, even if functionally similar (e.g. **Figure 3B;** label #2, **Supplementary Material 2)**. Any mask claiming multiple non-identical regulatory approvals is *prima facia* counterfeit. Several masks additionally including labels such as “PM 2.5” that are typically meant to denote protection from nuisance dust and air pollutants (label #6, **Supplementary Material 2)**. Such masks are likely fraudulently relabeled simply by stamping “N95” or “KN95” on the box.

**Figure 2:**
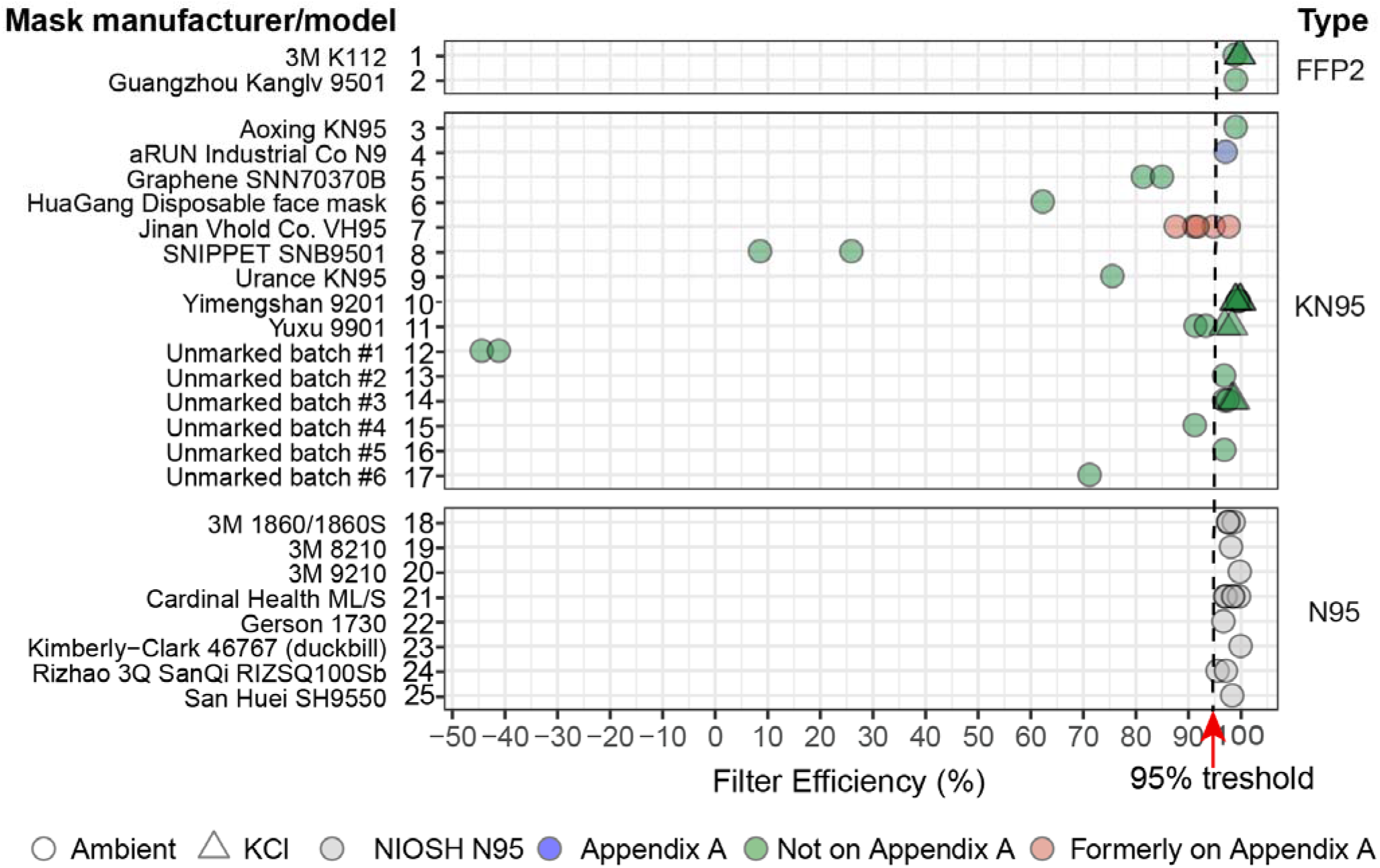
Filtration efficiency of N95-type masks using ambient particles and aerosolized KCl particles as testing agents. The lowest filtration efficiency recorded for any particle size tested in shown; full data are provided in Supplementary Table 3. Masks are grouped based on the testing standard they comply with (FFP2, KN95, or N95) but some masks incorrectly claim compliance with multiple standards. “NIOSH N95” refers to masks appearing on the list of *NIOSH-Approved N95 Particulate Filtering Facepiece Respirators* and regulated according to US standards; six of these models are manufactured in the US and the Rizhao and San Huei masks are manufactured in China; all of these masks were available on the US market prior to the current COVID pandemic. “Appendix A” refers to masks that are listed in the FDA EUA “*Non-NIOSH Approved Disposable Filtering Facepiece Respirators Manufactured in China*” first issuedon February 2020 and subsequently updated. These masks have been made available only as a result of the pandemic.

**Figure 3.**
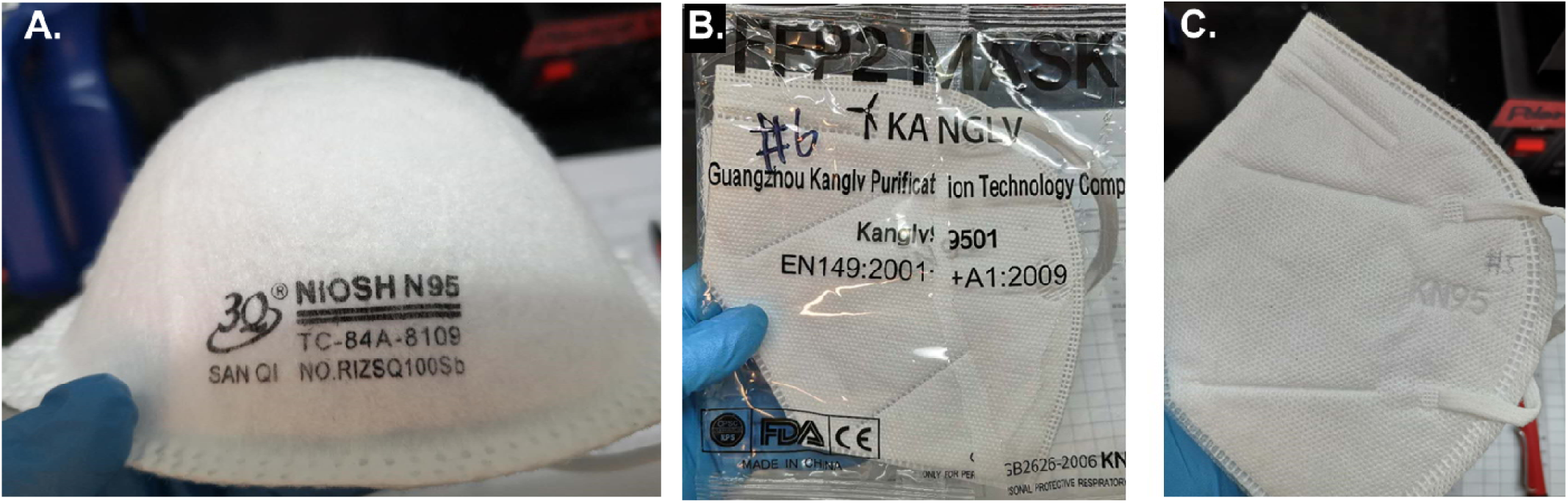
Images of a subset of masks subjected to performance testing and manufactured in China. **A**. A dome-type mask manufactured to N95 standards and listed on the NIOSH website for sale in the U.S. that has all of the required markings. This mask performed as expected (Figure 2, line 24). **B**. A flat-fold mask that claims compliance with European FFP2 but contains an FDA logo, which is not allowable. This mask performed well in our tests across all particle sizes and has the performance expected of a legitimate product (Figure 2, line 2). **C**. A flat-fold mask supplied in bulk with no markings other the embossed KN95 label; this mask had negative filtration efficiency, and more particles were detected at the output of our test apparatus than at the input (Figure 2, line 12). Additional photographs of mask are available in Supplementary Material 2.

After excluding brands that were visibly counterfeit based on these criteria, 18 of the most commonly donated mask models from the inventory were selected for further study. These masks had identifiable manufacturer markings and included two FFP2, nine KN95, and eight N95 respirators. In addition to the nine KN95 masks with markings, six unmarked KN95 masks that had been provided in bulk were selected for further investigation. N95 masks meeting NIOSH standards must have TC-approval numbers (11) printed on the mask and must be listed on the NIOSH Certified Equipment List (CEL) (16) or the NIOSH Trusted-Source list (17) (e.g. **Figure 3A**; label #24, **Supplementary Material 2**). NIOSH has an excellent infographic illustrating the correct labeling of N95 masks that we reproduce in supplementary materials for convenience (**Supplementary Material 1**). These CEL and Trusted-Source lists pre-date the COVID-19 pandemic and contain information on FFRs that would normally be available through traditional healthcare supply chains; several are manufactured in China. All eight N95 mask models evaluated had valid TC numbers and information could easily be obtained on them by searching the manufacturers’ websites. This is not necessarily a sufficient test for legitimacy because the CDC has reported that some counterfeiters steal TC-numbers from legitimate suppliers (11).

Masks claiming compliance with KN95 and FFR2 standards were cross-referenced with the FDA Appendix A list (13) and assessed for a valid business website associated with the brand. Neither of the FFP2 masks in our inventory are presently listed on Appendix A of the FDA EUA (13) (**Table 1**). Data on 3M K112 is readily available (18) and appears to be widely distributed in Europe but we found no reliable information on the Guangzhou Kanglv 9501 model (labels #1 and #2 respectively, **Supplementary Material 2**). Nine of the KN95 masks tested had markings on their surface or packaging and six were completely unmarked. Of the nine marked KN95 masks studied in detail, two were listed on Appendix A initially but one (Jinan VHOLD Co. VH95, label #7, **Supplementary Material 2**) was removed as of July 13, 2020 leaving only aRUN Industrial Co. N9 (Label #4); seven could not be matched to any brand or model on Appendix A based on information on the packaging or the mask itself (**Table 1;** labels #3 and 5, 6 and 8-11). Six additional KN95 mask types were completely unmarked and could therefore not be checked against Appendix A or a manufacturer’s website (**Figure 3C**; Labels #12-17, **Supplementary Material 2**).

### Testing mask performance

We subjected the masks described above to filtration performance testing at a university laboratory (**Figure 2**). Testing was performed on both ambient particulate matter and aerosolized potassium chloride (KCl) in the size range 0.3 to 10 μm, a relevant range for N95 FFRs. Passing this test is not sufficient to establish conformity with NIOSH, EN149 or GB2626 standards since all three involve a range of tests for multiple performance characteristics under carefully controlled conditions (3). However, we have found that results obtained using our testing system conform well to tests performed at a commercial pre-certification laboratory to NIOSH standards (see http://cleanmask.org/procedures for further details). Our testing revealed that all N95 masks and a subset of KN95 masks performed as expected in that they repeatedly removed >95% of particles down to 0.3 μm. However, a substantial number of KN95 models, both marked and unmarked, failed testing and one unmarked mask released more particles than were present at the input of the testing apparatus, which presents *negative* filtration performance (**Figure 2; Supplementary Material 3**). The single KN95 mask tested that is still listed on Appendix A exhibited greater than 95% filtration efficiency (aRUN Industrial Co. N9) whereas one mask model formerly on the FDA EUA Appendix A demonstrated less than 95% filtration efficiency (Jinan VHOLD Co., LTD Model VH95). These data confirm results from other sources, including the CDC (11) that poorly performing masks make up a substantial portion of the inventory of non-domestic N95 type masks available in major academic medical centers in the US. Fortunately, as of writing, none of the mask models analyzed in this paper saw use in clinical practice and they are currently being stored for potential emergency use.

Fit is a critical feature of N95-equivalent masks and is typically evaluated by end users using OSHA-regulated fit tests. It has been observed that KN95 masks with ear loops instead of headband straps often fail fit testing and this is a factor that must be considered when choosing a product (7,11,19). We have recently described devices for improving the fit of such masks using secondary mask frames (20). We observed that some masks labelled KN95 (a subset of the unmarked KN95s in **Figure 2**) have thin perforations and may also have embossed ‘KN95’ lettering that exposes the thin filter layer. This makes the masks particularly fragile and subject to ripping when donned. Such masks should probably be avoided.

## DISCUSSION

A growing number of investigators and Federal agencies have reported that many N95-equivalent masks manufactured overseas whose distribution in the US became possible due to recent FDA EUAs do not perform to relevant US and international standards (11). Our data show that, several months into the COVID-19 pandemic, these under-performing masks made up a substantial portion of the donated inventory at major medical centers in the US (**Figure 2**). Our performance testing, although limited in scope, suggests that some masks perform very poorly, removing only 8% - 80% of 0.3 μm particles; alarmingly, at least one mask added particulate matter to the airstream and therefore had negative efficiency. In many cases these masks claim compliance with multiple non-identical regulatory standards and they are clearly counterfeits. Remarkably, several models of KN95 that passed preliminary performance testing had little or no identifying markings, or had labelling that was inconsistent with listings in FDA EUA Appendix A: even legitimate KN95 masks lack vendor-specific information similar to the TC numbers required by NIOSH on all N95 masks. We devoted substantial effort to tracking down information on these KN95 masks but in many cases could not find corresponding manufacturers, distributors or informational websites. We conclude that is impossible in many cases to determine whether a KN95 mask is legitimate or not based on the label or packaging. Unfortunately, this would appear to be an ideal setting for counterfeiters.

### Recommendations for end users

Based on the current study, and in consultation with environmental and occupational health offices at three different hospitals, we propose the following guidelines for sourcing N95-equivalent masks:

1. **Use trusted supply chains**. Whenever possible, use trusted supply chains to provide products and ask for the technical datasheets or certification documents for a specific brand and model. These documents should not contain obvious spelling or grammatical errors. For all N95 and FFP2 that passed testing, these materials could easily be located on manufacturer websites.
2. **For FFRs claiming N95 certification**, check for active and correct TC numbers on the <u>NIOSH</u> <u>Certified Equipment List (CEL)</u> or the <u>NIOSH Trusted-Source</u>. Check that the TC number matches the style and manufacturer of the mask. Check that all other information matches NIOSH requirements (see infographic in **Supplementary Figure 1**).
3. **Check for similarity to a fraudulent product on the <u>NIOSH Web site</u>.** We recommend sending pictures of products falsely labelled as “N95” to the CDC so the agency can expand its on-line gallery and assist others in identifying products that should not be used under any circumstances. Even seemingly high-quality packaging can hide a nonfunctional product.
4. **For FFRs claiming compliance to a non-US standard (e**.**g. KN95s, FFP2s)**, check if masks are on the <u>FDA Appendix A</u> or <u>Exhibit 1</u> lists of respirators authorized for importation under EUA. Also check the <u>CDC Assessment Results for Not NIOSH-approved respirators</u> for filtration performance. If the mask is not on the list, it can be submitted for testing on the CDC <u>International Respirator Assessment Request</u> page.
5. **Check for inconsistent markings**. No FFP2/3, KN95, DS/DL, P2/3, or PFF product should bear a NIOSH stamp since NIOSH only certifies the US N95 standard (the reciprocal is also true). Additionally, fraudulent products often carry multiple labels (KN95, N95, FFP2). A list of different respirator certifications by nation is available at the CDC Website: https://www.cdc.gov/niosh/npptl/respirators/testing/NonNIOSH.html.
6. **Consider independently performing filtration testing** in the absence of verifiable manufacturer information for a specific mask. This can be accomplished by submitting the mask for testing to a CDC or a NIOSH-approved commercial facility. Some institutions may want to consider using their own testing apparatus, as described in the methods section of this paper and at http://cleanmask.org. Fit testing should be performed on all masks used in a healthcare setting.

For large donations of respirators from unknown suppliers, we recommend that quality assurance testing, including filtration testing, be performed before the respirators are issued to healthcare providers or other frontline personnel. Although we recognize that such testing is difficult to perform for many independent institutions, commercial pre-certification laboratories are able to provide this service at a reasonable cost and turnaround time. For example, the Manufacturing Emergency Response Team (<u>M-ERT</u>) has <u>collaborated with a network of local testing laboratories</u> across Massachusetts in response to the pandemic; their ability to provide functional testing of N95-type masks contributes to community resiliency. The possibility that counterfeit masks can have negative filtration efficiency strongly suggests that masks of unknown provenance, or masks whose manufacturer cannot be independently verified should not be used. High quality surgical masks are likely to be a safer option.

## CONCLUSIONS

The inconsistent and at times bewildering labeling on KN95 masks makes it difficult to identify manufacturers and determine if they are legitimate products. We recommend that all N95-type masks have identifying information printed directly on the product that identifies their manufacturer, such as numbers functionally similar to TC numbers for N95 masks. We also recommend that the FDA make public all data submitted by manufacturers listed in EUA Appendix A. All companies should be required to provide basic operational data including name and place of business, proprietary or brand name, model number, marketing authorization, a copy of the product labeling and evidence of authorization with quality management systems for healthcare devices (e.g. through 21 CFR Part 820, ISO 13485 or an equivalent) (21). Any legitimate company will have this information immediately available, although it may initially be provided in a foreign language. Such information is readily available for standard NIOSH-approved N95 masks, and this provides a template for Appendix A as well (e.g. a <u>listing of approved surgical N95</u> manufacturers and models that include links to legitimate corporate websites and donning instructions).

Since the initial EUA issuance, the FDA has twice amended information on non-NIOSH approved masks, once in May and once in June, to improve supply chain oversight. Additional criteria have been established for Appendix A listings including required CE marks (Conformitè Europëenne, denoting health, safety, and environmental protection standards for products sold within the European Economic Area(22)) or NMPA certification (National Medical Products Administration, a Chinese government agency for regulating pharmaceuticals, medical devices, and cosmetics (23)), to ascertain certification from a trusted notified body. The FDA and CDC have also initiated a large-scale testing program to randomly sample respirators imported from China and test their filtration ability, but this will be of limited use without a method for end users to link information on foreign-manufactured masks with test results, such as through use of TC and/or lot numbers. We also recommend stronger oversight of the respirator supply chain by Federal regulatory agencies, including required performance testing of non-NIOSH approved respirators prior to distribution, even in times of crisis. As the current pandemic evolves, generating and maintaining an updated list of trusted alternate suppliers will leave us better prepared for current and future supply shortages.

## Limitations of this study

The testing performed in this study uses readily available equipment but is not equivalent to NIOSH-approved testing. We have collected data on a set of N95 masks exposed to various sterilization procedures using the equipment described here and also using testing to NIOSH standards at a commercial laboratory (ICS Laboratories, USA; equipped to perform NIOSH pre-certification testing) (24). Instantaneous filtration efficiency values measured in the two tests for different masks of the same model undergoing the same sterilization procedure had a correlation coefficient of 0.89 and all masks demonstrating greater than 95% filtration efficiency also passed ICS tests (and vice versa; see cleanmask.org). We therefore conclude that our testing procedure provides a reasonable estimate of filtration performance for N95-type masks. Nonetheless, the results described here should be interpreted as relative, not absolute, measures of filtration efficiency and no mask should be considered suitable for human use based on our data alone.

## Data Availability

All data generated or analyzed during this study are included in this published article and its supplementary information files.

## LIST OF ABBREVIATIONS

CDC: Centers for Disease Control and Prevention
CEL: Certified Equipment List
EUA: Emergency Use Authorization
FDA: Food and Drug Administration
FFRs: Filtering Facepiece Respirator
NIOSH: National Institute for Occupational Safety and Health
PPE: Personal Protective Equipment

## DECLARATIONS

### Ethics approval and consent to participate

Not applicable.

### Consent for publication

Not applicable.

## Author Contributions

Assessing mask donations: S.H.Y., P.M., J.B.

Mask filtration testing: E.T, J.M., J.L.

Website construction: M.K.

Writing: D.P., E.T., A.K.C., H.Y., M.M.C, M.S.S., F.T.B., S.H.Y., N.R.L, J.L., P.K.S.

Greater Boston Pandemic Fabrication Team (PanFab) Consortium Coordination: D.P., H.Y., P.K.S.

## Acknowledgements

Above all we thank the members of the Greater Boston Pandemic Fabrication Team (PanFab) for technical, administrative, and logistic support necessary for the execution of this project. Membership found at https://www.panfab.org/the-team-and-the-project/consortium-members. Local fabricators, makers and citizens generously donated their time and resources and were essential for all stages of the project. This work was also supported by the Harvard MIT Center for Regulatory Sciences and by NIH/NCI grants U54-CA225088 (to PKS, NRL and DP) and by T32-GM007753 (to DP) and by the Harvard Ludwig Center.

## TABLES AND FIGURES

**Table 1:** Mask models donated to major academic medical centers in Boston during the COVID-19 pandemic and their corresponding regulatory designation. Highlighted models indicate masks models that underwent filtration testing at academic medical center. N95 model certification was checked in the NIOSH Certified Equipment List. *Known counterfeit masks are listed on CDC website; Suspected counterfeit masks were identified by guidance listed on the same website. ** Jinan VHOLD Co LTD VH95 was later removed from Appendix A.

## SUPPLEMENTARY MATERIALS

**Supplementary Material 1**: NIOSH infographic illustrating the correct labeling of N95 masks “NIOSH Diagram.pdf”.

**Supplementary Material 2:** Photographs of masks undergoing filtration efficiency testing “Photographs of masks tested.ppt”. Numbers on images correspond to number on Figure 2 and first column of Supplementary Material 3.

**Supplementary Material 3:** Filtration efficiency raw data (depicted in Figure 2) “Filtration Efficiency Data.csv”.

